# Associations between water supply intermittencies and drinking water quality, child health, and caregiver emotional stress in peri-urban Malawi

**DOI:** 10.1101/2025.06.03.25328821

**Authors:** Caitlin G Niven, Ben Clark, Emily Floess, Blessings Chirwa, Monica Matekenya, Emma Budden, Stella Cadono, John Chavula, Victor Chisamanga, Aubrey Dzinkambani, Chisomo Kaponda, Neema Ngondo, Norah Patterson, Sheena Symon, Brighton Austin Chunga, Rochelle H Holm, Petros Chigwechokha, Francis L. de los Reyes, Cassandra L Workman, Angela R Harris, Ayse Ercumen

## Abstract

**Background:** Achieving universal access to safe and affordable drinking water remains a critical global challenge, particularly in low- and middle-income countries where intermittencies in water supply are common. We aimed to examine relationships between water intermittency and multiple outcomes, including microbial water contamination, child health, and caregiver stress.

**Methods:** We conducted a cross-sectional study with 237 households with a child <5 years old in a peri-urban neighborhood of Blantyre, Malawi. We conducted a structured questionnaire with primary caregivers to record water intermittencies, water handling and hygiene practices, caregiver-reported child diarrhea and acute respiratory infection (ARI) symptoms, and stress among caregivers. Drinking water samples were tested for *E. coli* and cefotaxime-resistant *E. coli* using IDEXX Quanti-Tray/2000. We used generalized linear models to evaluate how intermittency occurrence, frequency, and duration influenced outcomes, adjusting for sociodemographic and WASH factors.

**Findings:** Of 237 households, 32.5% reported ≥1 water intermittency in the past month. These households were more likely to experience water insecurity, skip bathing and laundry, and report less handwashing after animal contact or outdoor work. *E. coli* was detected in 65.7% and cefotaxime-resistant *E. coli* in 8.4% of water samples. Intermittency was not associated with impaired water quality. Children in intermittent households had higher prevalence of diarrhea (PR=1.94, 95% CI: 1.11–3.39) and ARI with fever (PR=2.00, 95% CI: 1.11–3.60). Rare/short intermittencies were more strongly associated with diarrhea; frequent/long intermittencies were more strongly associated with ARI. Caregivers in households with short and frequent intermittencies reported higher stress.

**Interpretation:** Water intermittencies were associated with impaired hygiene, child illness, and caregiver stress. Frequency and duration modified the associations, suggesting that short- vs. long-term behavioral adaptations may differently influence exposure to enteric and respiratory pathogens and stress responses. Interventions like low-flow handwashing stations and water reuse may help reduce health risks in intermittent water settings.

## 1. Introduction

The United Nation’s Sustainable Development Goal 6.1 aims to achieve universal access to safe and affordable drinking water by 2030; however, meeting this goal remains a significant global challenge. The WHO/UNICEF Joint Monitoring Programme (JMP) estimated that, as of 2022, 2.2 billion individuals lacked access to safely managed drinking water (United Nations Children’s Fund (UNICEF) & World Health Organization (WHO), 2023). A recent study using household-level estimates from geospatial earth observation data rather than national-scale assessments estimated that up to 4 billion individuals in low- and middle-income countries do not have access to safely managed drinking water at home (Greenwood et al., 2024). The JMP defines “safely managed” water service as water from an improved source that is located on premises, free of fecal and priority chemical contamination, and available when needed (United Nations Children’s Fund (UNICEF) & World Health Organization (WHO), 2023); for piped water supplies, they define “available when needed” as 12 or more hours of water service per day. The WHO emphasizes that an improved water source must not only be safe but also continuously available on the premises (WHO, 2023). However, intermittent piped supplies serve over one billion individuals, approximately 12.5% of the global population (Ferrante et al., 2024), and non-piped supplies such as boreholes are subject to prolonged interruptions due to wellhead malfunction, water scarcity or both (MacAllister et al., 2022).

Lack of continuous water access has profound implications for water security and quality, hygiene practices, health, and emotional well-being. Water insecurity at the household level refers to the inability to access adequate, reliable, and safe water for everyday use, including water for drinking, washing, cooking, and cleaning (Jepson et al., 2017; Young et al., 2019). For piped supplies, intermittencies in service allow pathogens to intrude into pipes during non-pressurized periods, compromising water quality. Microbiological contamination levels can be up to 45 times higher in intermittently supplied taps, compared to continuously supplied taps (Kumpel & Nelson, 2013). Intermittent or unreliable water supplies can also force households to store water for extended periods, ration water usage, or supplement their primary supply with water from unsafe/unprotected sources (Cronk et al., 2024). Opportunities for contamination at various points during water collection and storage increase the risk of waterborne diseases. These contamination events may also foster the proliferation of antimicrobial-resistant bacteria in drinking water due to contamination with human or animal waste in stagnant pipes and/or under suboptimal water storage conditions (Burnham, 2021; Hayward et al., 2020; Yu et al., 2022). Though antimicrobial resistance has been evaluated extensively in aquatic environments, few studies to date have assessed the effect of water intermittencies on contamination of drinking water with antimicrobial-resistant bacteria and subsequent proliferation and dissemination (Kumpel & Nelson, 2016; Rayasam et al., 2019; Taviani et al., 2022; Tokajian & Hashwa, 2004).

Further, water intermittency and limited water availability can contribute to the spread of water-washed diseases by impeding on essential hygiene practices, as households may have to prioritize the use of clean water for other household tasks (United Nations Children’s Fund (UNICEF) & World Health Organization (WHO), 2020). This potentially limits hygiene practices such as handwashing, which has been linked to higher rates of enteric and acute respiratory infections (ARIs) (Ashraf et al., 2020; Freeman et al., 2014; Ross et al., 2023; Swarthout et al., 2020). Intermittencies in water service have been associated with increased risk of gastrointestinal diseases like diarrhea, typhoid fever and enteric infections (Bivins et al., 2017; Ercumen et al., 2015) but studies specifically examining their potential effect on respiratory infections remain scarce (Parisoto et al., 2024; Ray, 2020). Diarrhea and ARIs are among the major drivers of antibiotic use (Hassan et al., 2021; Rhee et al., 2019; Tekleab et al., 2017), therefore, increased enteric and respiratory infections associated with intermittent water supplies can plausibly lead to increased antibiotic use and consequently increase community carriage of antimicrobial resistance.

The direct and indirect effects of lacking continuous access to water extend beyond physical health outcomes, and can significantly impact mental and emotional health and wellbeing through increased psychosocial stress and emotional burdens (Jepson et al., 2017; Smiley & Stoler, 2020, Thomson 2024). Daily challenges in finding an adequate water supply, ensuring sufficient water is stored, and reserving or rationing water for priority purposes can contribute to chronic stress and anxiety (Collins et al., 2019; O’Brien et al., 2024; Stevenson et al., 2012; Thomson et al., 2024; Workman et al., 2024). Women and children, who are primarily responsible for water-related chores, particularly in sub-Saharan Africa, often bear the greatest emotional and physical burden (Rhue et al., 2023; Stevenson et al., 2012; Workman & Ureksoy, 2017). Studies in sub-Saharan Africa have found women must balance responsibilities with the burden of water-related chores such as fetching, treating, boiling, storing, etc. (Adams, 2024; Brewis et al., 2024). The physically demanding task of fetching and managing water can contribute to significant stress, especially when caregivers must make difficult decisions about prioritizing limited water for essential household needs. Chronic stress, anxiety, and a sense of helplessness are common among those facing uncertainty about access to safe and sufficient water (Collins et al., 2019; O’Brien et al., 2024; Stevenson et al., 2012; Workman & Ureksoy, 2017; Wutich et al., 2020). A study in Ethiopia found that up to 60% of caregivers reported anxiety and social conflict related to water scarcity, highlighting the widespread emotional toll (Stevenson et al., 2012). Moreover, water insecurity does not exist in isolation, it is compounded by other stressors such as lack of access to safe sanitation, increased risks of waterborne diseases, conflicts over scarce resources and food insecurity (Brewis et al., 2020; Workman et al., 2022). The cumulative impact of these challenges – heightened disease risks, physical demands, and emotional strain – demonstrates how water insecurity, including intermittent water access, is both a physical threat and a significant psychosocial burden.

Malawi is a landlocked country in Southern Africa with a population of over 20 million people, ranking 172 out of 193 on the Human Development Index, placing it among the poorest 8% of countries globally (United Nations, 2025). More than half the population lives in poverty and one-fifth in extreme poverty (World Bank, 2024), with over a quarter unable to afford the recommended daily food intake—putting many individuals at risk for elevated stress and adverse health outcomes (Coulibaly et al., 2015; McCarthy et al., 2021). Water insecurity is also widespread: 3.8 million Malawians lack access to clean drinking water, with rural access (65%) lagging behind urban areas (90%) (Coulibaly et al., 2015). Previous studies in Malawi have highlighted several challenges with water services. For example, piped water systems face sustainability issues due to logistical, technical, and infrastructural constraints (Holm et al., 2022). In informal settlements in Lilongwe, the capital city, limited and unreliable water kiosks force households to endure long queues and service gaps (Adams, 2018). At the utility level, water boards struggle with high levels of unaccounted-for water, mainly from leaks, undermining supply reliability (Harawa et al., 2016; Kalulu & Hoko, 2010).

This study aims to examine the relationship between intermittencies in water supplies and a comprehensive range of outcomes, including drinking water contamination with fecal indicators and antimicrobial-resistant bacteria, child health outcomes (diarrhea, ARI, and antibiotic use), and caregiver emotional stress, within peri-urban Malawian households. While previous research has individually explored associations between water intermittency and some of these outcomes, this study builds on existing literature by investigating multiple outcomes along the causal chain to corroborate mechanisms.

## 2. Methods

### 2.1 Study Design and Population

We systematically enrolled 237 households in Bangwe, a peri-urban town in Blantyre, Malawi. We used GIS to select random starting points within the town boundaries and systematically approached every third household in each direction from the starting point to ensure representative geographic coverage. Households were eligible for enrollment if they had at least one child under the age of five and if an adult over the age of eighteen that was responsible for those children was available and consented to participate. Enrolled households were visited once to conduct a structured questionnaire and collect samples of drinking water.

### 2.2 Data Collection

Trained enumerators from Mzuzu University (MZUNI) and Malawi University of Science and Technology (MUST) administered a structured questionnaire among the primary caregivers in the enrolled households to record water intermittencies, child health outcomes and antibiotic use, perceived respondent stress, as well as relevant demographic information and water, sanitation and hygiene indicators. Water intermittency was enumerated using the 12-item Household Water InSecurity Experiences (HWISE) scale that assesses the frequency of experiences related to water insecurity, including unavailability, intermittency, and associated negative emotional responses, using a five-point Likert scale ranging from never (0) to always (20+ times) in the last four weeks (Young et al., 2019). One of the 12 HWISE items is specifically focused on intermittency, asking how often the household’s primary water source has been interrupted or limited in the previous four weeks. In addition, we recorded the duration of the intermittency by asking an additional question on how long the last interruption lasted before water service was restored. Recorded child health outcomes included the reported occurrence of diarrhea (as defined by caregiver), loose stools, blood in stool, constant cough, congestion/runny nose, difficulty breathing, and fever within the last seven days. We also recorded the reported occurrence of two negative control outcomes (outcomes theoretically not associated with water intermittency) (Arnold & Ercumen, 2016; Lipsitch et al., 2010), including ear infection and a rash anywhere on the child’s body within the last seven days. Respondent stress was recorded using the 10-item Perceived Stress Scale (PSS) that evaluates feelings and thoughts in the past month as they pertain to how unpredictable, uncontrollable or overwhelming events are in life using a five-point Likert scale ranging from never (0) to very often (4) (Cohen et al., 1983; Harris et al., 2023).

### 2.3 Sample Collection and Processing

Enumerators trained in aseptic technique collected drinking water samples from enrolled households. Enumerators asked the respondent for a glass of drinking water in the same way they would give it to a child under the age of five. Approximately 200 mL of sample was transferred into a sterile WhirlPak bag that contained sodium thiosulfate to remove any residual chlorine (Whirl-Pak Filtration Group, USA). The enumerators requested an additional glass of water if the first glass contained less than 200 mL water. The enumerators recorded whether the sample was obtained from a storage container, the source of the water and whether it had been treated to make it safer for consumption. Field blanks were collected by placing a WhirlPak bag pre-filled with 100 mL sterile DI water into the field cooler of a randomly selected enumerator and asking them to return it to the laboratory along with the samples. One field blank was collected for approximately every 30 water samples (3.0%, 7/236). Samples were transported on ice to the laboratory at MUST and processed within five hours.

We used IDEXX Quanti-Tray/2000 with Colilert-18 media (IDEXX Laboratories, Westbrook, ME, USA) with and without antibiotic supplementation to enumerate the most probable number (MPN) of *E. coli* and cefotaxime-resistant *E. coli*. Cefotaxime-resistant *E. coli* was used as a presumptive indicator for extended-spectrum beta-lactamase (ESBL)-producing *E. coli*, which are of particular concern due to their resistance to broad-spectrum beta-lactam antibiotics and form the basis of the WHO’S Tricycle global surveillance initiative (WHO, 2021). 100 mL of sample was added to a sterile WhirlPak, to which Colilert was added and gently dissolved before dispensing the sample into a Quanti-Tray to be sealed. A second 100 mL aliquot from the same sample was analyzed using the same process plus the addition of 80 µL of 5 mg/mL filter-sterilized cefotaxime solution (Hornsby et al., 2023). Lab blanks were analyzed using the same steps with 100 mL of sterile DI water. One lab blank was processed for approximately every 10 water samples (11.9%, 28/236). Following incubation for 18 hours between 35-36°C, Quanti-Trays were read by counting the number of wells that turned yellow, indicating the presence of total coliforms, and those that were both yellow and fluoresced under long-wave UV light, indicating the presence of *E. coli*.

### 2.4 Ethics

The research protocol was approved by the Research Ethics Committee (MZUNIREC) at Mzuzu University (MZUNIREC/DOR/24/38) and University of North Carolina, Greensboro (UNC-Greensboro IRB #: 20-0245), with additional ethical oversight from North Carolina State University via Inter-Institutional Agreement. All participants provided written informed consent in the local language (Chichewa).

### 2.5 Statistical Analysis

#### Exposure variables

We classified water intermittency in three different ways based on the survey responses: occurrence, frequency, and duration of intermittency. We used the HWISE intermittency question to generate a binary variable for whether the household experienced any intermittencies vs. no intermittency in their water service in the last four weeks. Based on the same question, we also generated a categorical variable for the frequency of intermittencies, including none, rarely (1-2 times), and sometimes/often/always (3-20+ times). We generated a categorical variable for the duration of the last intermittency the household experienced in the last four weeks, including none, below-median duration, and above-median duration.

#### Outcome variables

Outcomes were evaluated across the causal chain, spanning water quality, child health and caregiver stress (Fig 1). Water quality outcomes included the prevalence and log-10 transformed most probable number (MPN) of generic *E. coli* and cefotaxime-resistant *E. coli*. We replaced non-detects with half the detection limit (0.5 MPN/100 mL) and values above the detection limit of 2419.6 MPN/100 mL with 2420 MPN/100 mL before taking the logarithm. For samples that were positive for cefotaxime-resistant *E. coli*, we calculated the relative abundance as the ratio of cefotaxime-resistant *E. coli* counts to generic *E. coli* counts for the same sample and reported the resulting range as percentages. Child health outcomes were evaluated as the caregiver-reported 7-day prevalence of diarrhea, ARIs, and ARIs with fever as well as the 7-day prevalence of rashes and ear infections (our negative control outcomes). We used two different diarrhea definitions (Pickering et al., 2018). Caregiver-defined diarrhea recorded the caregivers’ response to the question whether their child had diarrhea, capturing their personal perception of what would constitute diarrhea or illness in their child. WHO-defined diarrhea was classified as three or more loose or liquid stools within a 24-hour period, based on the caregiver-reported symptoms. ARI was defined as a constant cough coupled with panting, wheezing, or having difficulty breathing; ARI with fever included the same symptoms with a fever occurring in parallel. Additional child health outcomes were whether the child took antibiotics and the number of times the child took antibiotics in the previous four weeks. We generated two composite variables for caregiver stress. We summed the scores to the individual Likert-scale questions to calculate a PSS score, and we generated a binary variable to define high stress (PSS score ≥27 out of 40) (Pieh et al., 2022). Additionally, we generated a binary version of each individual Likert-scale stress indicator by grouping responses of never, rarely and sometimes into a low-stress category and responses of often and always into a high-stress category for each individual indicator (Hanif et al., 2024).

**Fig 1.**
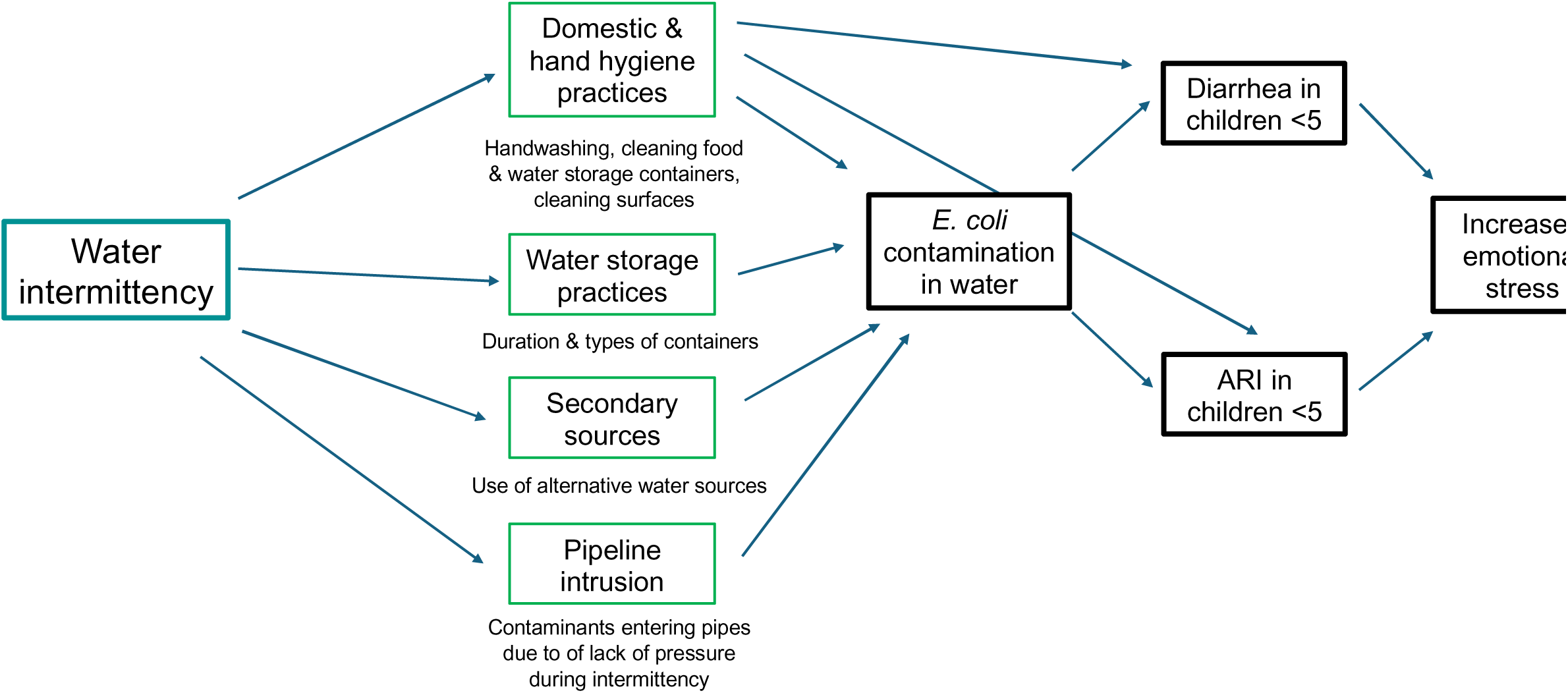
Directed acyclic graph (DAG) illustrating the hypothesized causal pathway between water intermittencies (in teal), mediating behaviors (in green) and our measured outcomes (in black) of water quality, child health, and respondent emotional stress.

#### Analysis approach

We used generalized linear models to assess the relationship between the different water intermittency definitions and each outcome variable. We used a Poisson error distribution with a log link for binary outcomes, negative binomial error distribution with a log link for over-dispersed count outcomes, a Gaussian error distribution with an identity link for continuous outcomes. Each model was adjusted for relevant covariates to control for potential confounding, including sociodemographic factors (e.g., household wealth quintile, education level, floor material), water, sanitation and hygiene indicators (e.g., use of an improved latrine and improved drinking water source according to JMP definitions, presence of a handwashing station with soap), and animal ownership (Text S1). We did not control for covariates on water use and hygiene practices as we hypothesized these to be on the causal pathway (Fig 1). We excluded binary and categorical confounders that did not sufficiently vary across our sample (<5% prevalence in any stratum). Models implemented robust standard errors to account for clustered outcomes. A cluster was defined as a geographic grouping of households that is at least 100 meters distant from the next group of clustered households (*Find Point Clusters—ArcGIS Online | Documentation*, n.d.). We used ArcGIS (ESRI, Redlands, California) to define the clusters using the defined distance (DBBSCAN) method based on GPS coordinates of enrolled households. Statistical analyses were conducted in R (version 4.3.1, RStudio (2023.06.0+421).

## 3. Results

### Participant characteristics

We enrolled 237 households between June 20-July 11, 2024. Study households were located within 38 unique spatial clusters based on their GPS coordinates. The mean age of respondents was 31.1 years, and households had on average 1.2 children under the age of five years old (Table 1). Approximately 40% of respondents had some primary education and 50% had some secondary education. On average, households spent $15-18 USD on weekly expenditures. Almost all households had floors made of cement/concrete, walls made of brick and roofs made of tin. The most common fuel type was charcoal, and the most common stove type was traditional solid fuel stoves (Table 1). Almost all households reported having a latrine, and 65.4% of households had an improved latrine; the most common latrine type was a twin pit latrine with a slab (Table 1). The primary source of drinking water was an improved source for 84.4% of households, including piped supplies (in own dwelling, own yard/plot or outside the compound) (56.6%) and boreholes (21.5%) (Table 1).

**Table 1.**
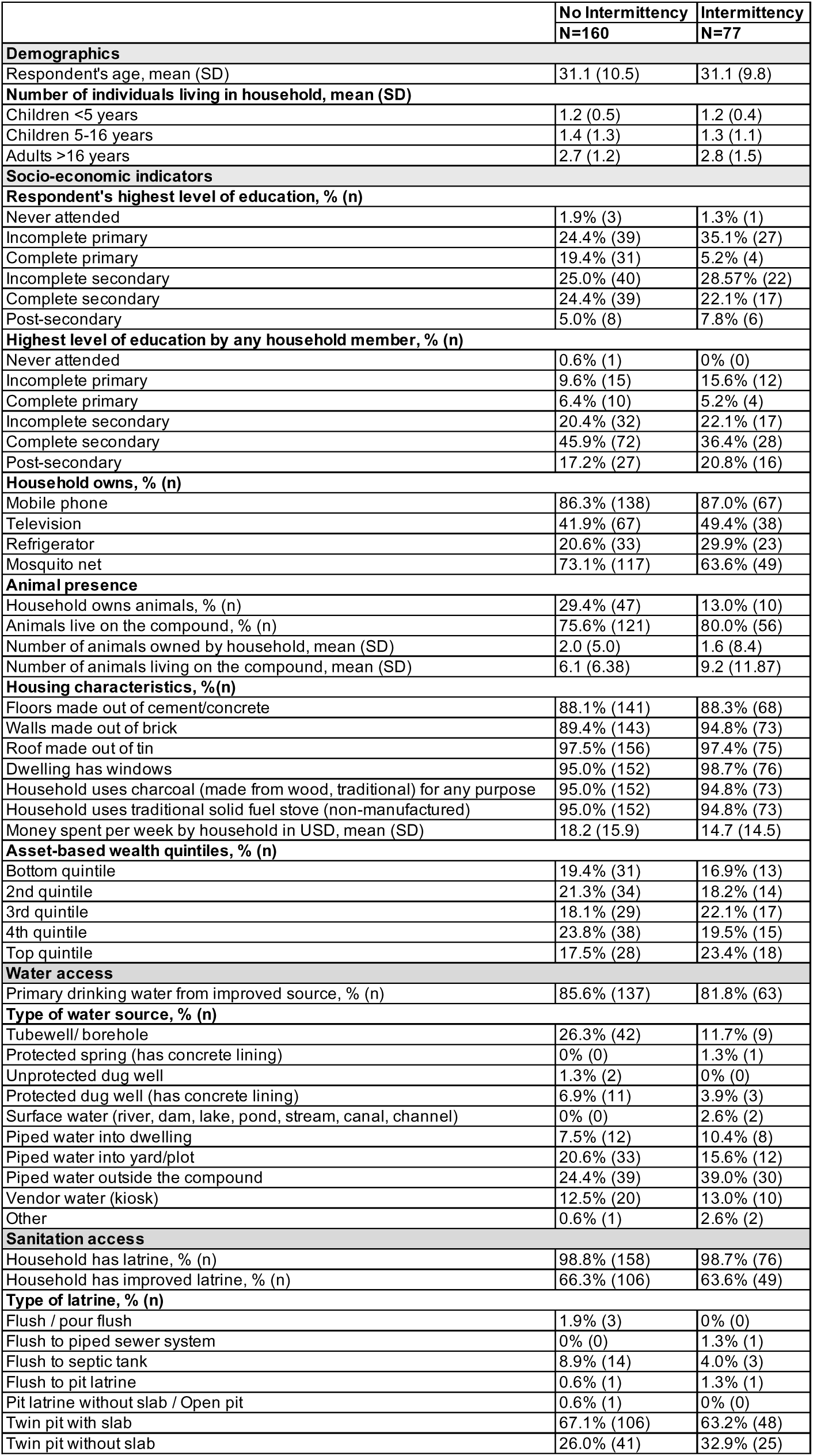
Demographic, socioeconomic and water and sanitation indicators among households experiencing at least one water intermittency vs. no water intermittency in the last four weeks.

#### Water intermittencies

Of 237 households, approximately one third (32.5%, 77/237) reported experiencing at least one water intermittency in the last four weeks. Specifically, 15.2% (36) of households experienced an intermittency rarely (1– 2 times), 12.2% sometimes (3–10 times), 3.8% often (11–20 times) and 1.3% always (>20 times). The mean duration of water intermittency was 4.53 days, and the median duration was 1.27 days (range: 0.02–90.31 days). Among the 77 households who experienced a water intermittency, 13.0% (10) of households were notified of the intermittency in advance, 2.6% (2) could predict the intermittency based on season or previous pattern of intermittencies, and 84.4% (64) were unaware of the upcoming intermittency.

Households with vs. without water intermittencies were similar in their overall characteristics (Table 1). However, households that experienced intermittencies were more likely to obtain their primary drinking water from a piped connection outside the compound (39.0% vs. 24.4%) and less likely to obtain it from a borehole (11.7% vs. 26.3%) (Table 1). Additionally, households with intermittencies appeared less likely to have at least one household member with complete secondary education (36.4% vs. 45.9%), to own a mosquito net (63.6% vs. 73.1%) and to own animals (13.0% vs. 29.4%) but more likely to own a television (49.4% vs. 41.9%) and a fridge (29.9% vs. 20.6%) and to report animals living on their compound (80.0% vs. 75.6%) than households that experienced no intermittencies (Table 1).

#### Water-related behaviors

*Domestic and personal hygiene*. Households with vs. without water intermittencies reported similar access to water for handwashing; approximately 10% households in both groups had a designated handwashing station and 9% had a handwashing station with available water observed at the time of the enumerators’ visit (Table 2). Intermittent households had higher mean household water insecurity scores than non-intermittent households (8.1 vs. 1.9 out of a maximum possible score of 36), and were more likely to be classified as water-insecure (HWISE score ≥12) (20.8% (16) vs. 4.4% (7) of households). Removing the water intermittency question from the total HWISE score reduced the mean score for intermittent households to 6.4 but did not change the score for non-intermittent households (Table 2). Using the truncated HWISE score that excludes the intermittency question, households with frequent intermittencies had higher mean HWISE scores than those with rare intermittencies (7.1 vs. 5.6), and households with longer intermittencies had similar mean scores as those with shorter intermittencies (6.2 vs. 6.6) (Table S1).

**Table 2.** Hygiene practices, water storage and secondary water sources among households experiencing at least one water intermittency vs. no water intermittency in the last four weeks.

|  | No Intermittency<br>N=160 | Intermittency<br>N=77 |
| --- | --- | --- |
| **Domestic and hand hygiene** |  |  |
| Household has handwashing station, % (n) | 9.4% (15) | 11.7% (9) |
| Household has handwashing station with water, % (n) | 8.8% (14) | 9.1% (7) |
| Household water insecurity score, mean (SD) | 1.9 (4.2) | 8.1 (5.9) |
| Truncated household water insecurity score, mean (SD) a | 1.9 (4.2) | 6.4 (5.8) |
| **Had to go without sometimes/often/always, % (n)** |  |  |
| Washing clothes | 9.4% (15) | 36.4% (28) |
| Bathing | 1.3% (2) | 10.4% (8) |
| Washing hands after dirty activities | 3.1% (5) | 5.2% (4) |
| **Reported handwashing occasions, % (n)** |  |  |
| Never | 1.3% (2) | 0.0% (0) |
| After defecation | 90.0% (144) | 94.8% (73) |
| After handling child's waste | 63.1% (101) | 61.0% (47) |
| After handling domestic animals | 10.0% (16) | 3.9% (3) |
| After handling animal feces | 8.8% (14) | 3.9% (3) |
| After working (garden, market, etc.) | 25.0% (40) | 15.6% (12) |
| After eating | 81.3% (130) | 79.2% (61) |
| Before eating | 91.9% (147) | 94.8% (73) |
| Before preparing food | 69.4% (111) | 62.3% (48) |
| Before feeding child | 41.3% (66) | 42.9% (33) |
| Before handling water (storage) | 23.8% (38) | 24.7% (19) |
| Before breastfeeding | 7.5% (12) | 9.1% (7) |
| **Water storage** |  |  |
| Water sample provided from storage container, % (n) | 87.5% (140) | 96.1% (74) |
| Stored water container fully or partially covered, % (n) | 78.8% (126) | 80.5% (62) |
| Stored water container has narrow mouth, % (n) | 17.5% (28) | 5.2% (4) |
| Duration of water storage in hours, mean (SD, range) | 32.6 (54.8, 0-577) | 29.9 (37.3, 0-194) |
| Water sample reported to be treated, % (n) | 5.6% (9) | 2.6% (2) |
| **Secondary water sources** |  |  |
| Primary drinking water source feels acceptable, % (n) | 89.4% (143) | 84.4% (65) |
| **Household obtains, % (n)** |  |  |
| Drinking water from secondary source | 24.4% (39) | 32.5% (25) |
| Drinking water from improved secondary source | 8.8% (14) | 18.2% (14) |
| Non-drinking water from improved source | 65.6% (105) | 64.9% (50) |

Water insecurity was also reflected in specific domestic hygiene behaviors reported by respondents in response to individual HWISE questions. Households with intermittencies were substantially more likely to go without doing laundry (36.4% vs. 9.4%) and bathing (10.4% vs. 1.3%) and slightly more likely to go without handwashing after dirty activities, such as defecating or changing diapers and cleaning animal dung (5.2% vs. 3.1%), sometimes, often, or always (3 to 20+ times) in the last 4 weeks (Table 2). When asked an unprompted question about when they wash their hands, respondents in both groups reported similar handwashing prevalence after defecating or handling child feces, and before eating or handling food or water (Table 2). However, intermittent households were less likely to report handwashing after handling animals (3.9% vs. 10.0%), handling animal feces (3.9% vs. 8.8%), or working outside (e.g. garden, market) (15.6% vs. 25.0%) than non-intermittent households (Table 2).

Households with frequent intermittencies were more likely to go without doing laundry, bathing and handwashing after dirty activities than households with rare intermittencies (Table S1). However, they were more likely to report handwashing after handling children’s waste, before feeding child, before breastfeeding and before handling water than those with rare intermittencies. Households with long intermittencies were less likely to report handwashing after handling domestic animals, handling animal feces, after working, before handling water, and before breastfeeding (Table S1). However, they were more likely to have a handwashing station with water compared to both households with short or no intermittencies.

#### Water storage, handling and use of secondary sources

A higher percentage of intermittent households provided drinking water samples from storage containers than non-intermittent households (96.1% vs. 87.5%) (Table 2). Approximately 80% of storage containers in both groups were observed to be covered but intermittent households were less likely to use narrow-mouthed containers (5.2% vs. 17.5%). The mean duration of water storage was slightly shorter in intermittent households compared to non-intermittent households (29.9 vs. 32.6 hours) (Table 2). The reported prevalence of treating drinking water in the home was low in both groups, but lower in intermittent households (2.6% vs. 5.6%). Intermittent households were also somewhat less likely to rate their primary drinking water source as acceptable (84.4% vs. 89.4%) and more likely to use secondary drinking water sources (32.5% vs. 24.4%) (Table 2). Households with rare and short intermittencies were more likely to provide drinking water samples from storage containers, have longer storage duration and obtain drinking water from secondary sources than those with frequent or long intermittencies (Table S1).

### Water quality

Of 236 drinking water samples, 65.7% (155) contained *E. coli* and the mean *E. coli* count was 0.74 log10-MPN/100 mL. Of 226 samples processed for with cefotaxime supplementation, 8.4% (19) contained cefotaxime-resistant *E. coli* and the mean count was −0.23 log10-MPN/100 mL. Among the 19 positive samples, the relative abundance of cefotaxime resistance (ratio of cefotaxime-resistant *E. coli* counts to generic *E. coli* counts for the same sample), ranged from 0.1% to 70%. Separated according to water source, samples with the highest levels of mean contamination ranged from 866.4 log10-MPN/100 mL for protected spring (n=1), 217.3 log10-MPN/100 mL for boreholes (n=51), 135.6 log10-MPN/100 mL for piped water into the dwelling (n=20), and 81.3 log10-MPN/100 mL for piped water outside the compound (n=69) (Table S2).

In adjusted analyses, intermittent and non-intermittent households had similar prevalence and counts of both generic and cefotaxime-resistant *E. coli* (Table S3). Water quality outcomes also did not differ significantly by intermittency frequency or duration. There were no significant differences in *E. coli* prevalence or counts between households that experienced intermittencies rarely (1-2 times), often/sometimes/always (3 to 20+ times) versus never (Table S4) or between households that experienced intermittencies of above-median duration (≥1.27 days), below-median duration (<1.27 days) versus never (Table S5). We could not assess associations between these categorical exposure variables and the prevalence or counts of cefotaxime-resistant *E. coli* because of the small number of positive samples.

### Child health

Among children in non-intermittent households, the 7-day prevalence of health outcomes was 16.3% for caregiver-defined diarrhea, 13.9% for WHO-defined diarrhea, 40.6% for ARI and 17.3% for ARI with fever (Table S6). In this group, 29.7% of children were reported to have taken antibiotics at least once in the last four weeks (Table S6). Children in intermittent households had about 60-90% higher prevalence of diarrheal illness, both for caregiver-defined diarrhea (prevalence ratio [PR]= 1.94 (1.11–3.39), p=0.02) and WHO-defined diarrhea (PR=1.63, (0.78–3.39), p=0.19) but the association for WHO-defined diarrhea could not be distinguished from chance (Table S6, Fig 2). Children in households with an intermittency also had 2 times higher prevalence of ARI with fever (PR=2.00, (1.11–3.60), p=0.02) and though borderline significant, 43% higher prevalence of ARI (PR=1.43 (0.99–2.09), p=0.06) (Table S6, Fig 2). There were no significant associations between water intermittency and child antibiotic use (Table S6, Fig 2). Among our negative control outcomes, water intermittency was not associated with the prevalence of having a rash (PR=1.25, (0.60-2.62), p=0.55) (Table S6, Fig 2). We could not assess associations for our second negative control outcome, because only 2.4% (7) of children were reported to have ear infections.

**Fig 2.**
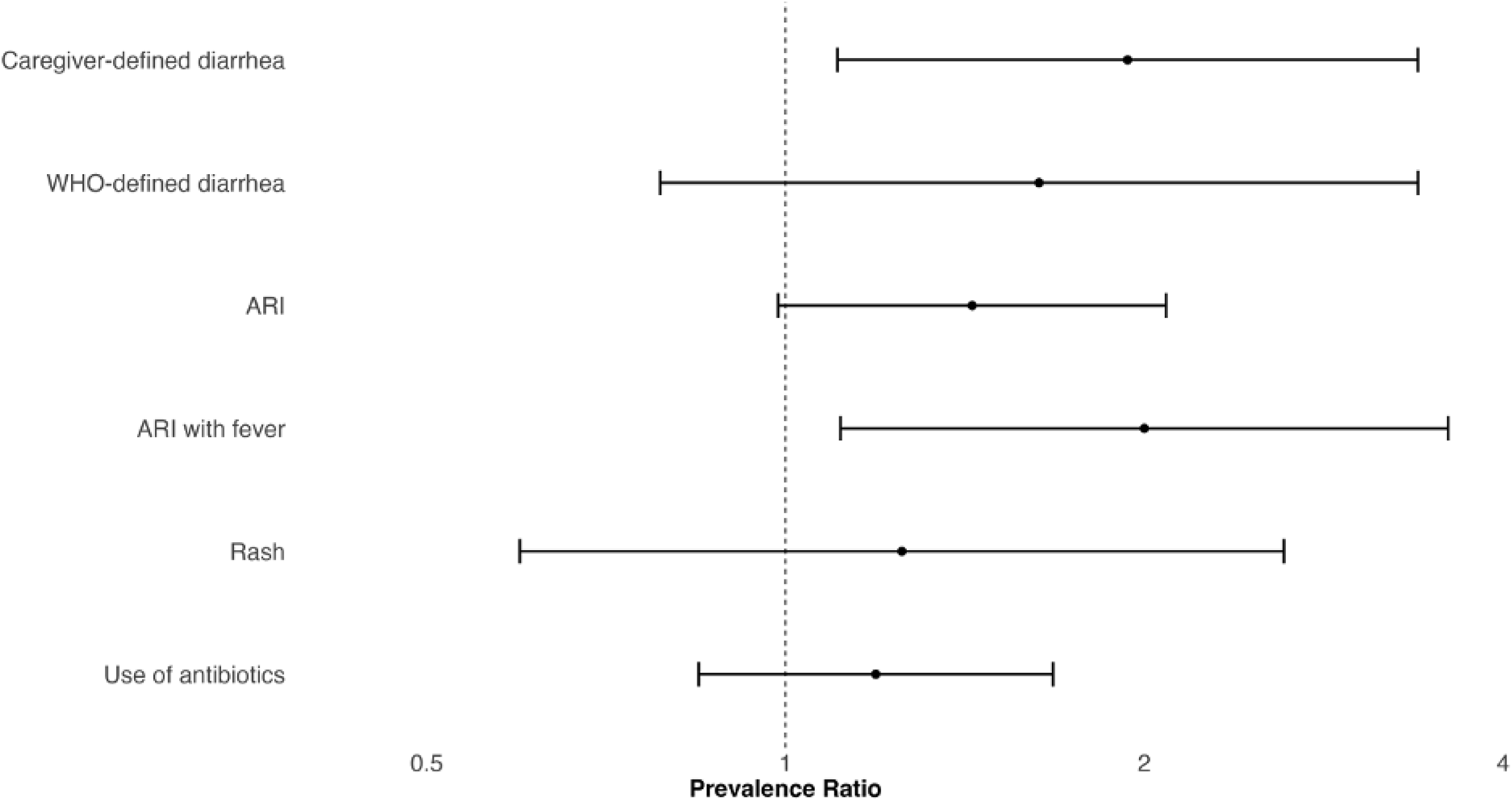
Forest plot of adjusted associations between water intermittency and caregiver-reported child health outcomes. The circles denote point estimates for prevalence ratios and the horizontal lines denote 95% confidence intervals. Models controlled for age of the respondent, age of the target child, total number of individuals living in household, respondent’s highest level of education, highest level of education level for anyone in household, asset-based household wealth quintile, average weekly expenditures by household household food insecurity score, whether household used piped water for their primary water source, whether household’s designated handwashing station had soap, whether household had an improved latrine (as defined by the WHO), flooring material inside household, total number of animals in compound, and respondent’s report of the last time it rained.

When analyzed by categories of intermittency frequency, children in households experiencing intermittency rarely had approximately 2 times higher prevalence of caregiver-defined diarrhea and WHO-defined diarrhea and used antibiotics twice as many times compared to children in non-intermittent households but the association for WHO-defined diarrhea could not be distinguished from chance (Table S7). Children in households experiencing intermittency sometimes, often, or always had between 1.70–2.3 times higher prevalence of ARI and ARI with fever than those in non-intermittent households; there were no associations with other health outcomes (Table S7).

When analyzed by categories of intermittency duration, children in households experiencing an intermittency of below-median duration had a significantly higher prevalence of caregiver-defined diarrhea compared to those with no intermittency (PR=2.21 (1.05–4.65), p=0.04) (Table S8). They also appeared to have higher prevalence of WHO-defined diarrhea and a higher number of antibiotic use episodes, but these associations could not be distinguished from chance (Table S8). Children in households experiencing an intermittency of above-median duration had significantly higher prevalence of ARI (PR=1.61 (1.02–2.55), p=0.04) and ARI with fever (PR=2.58 (1.53–4.34), p<0.0005) compared to those in non-intermittent households; there were no associations with other health outcomes (Table S8).

### Caregiver stress

Caregivers in households with water intermittencies had higher PSS composite scores (21.6 vs. 18.8 out of a maximum possible score of 40) and higher probability of experiencing high stress (PSS score ≥27) (22.4% vs. 9.4%) but associations could not be distinguished from chance after adjusting for confounders (Table S9, Fig 3). Caregivers in intermittent households were less likely to report feeling unable to control irritations (PR=0.36 (0.19, 0.71), p=0.003) but more likely to report feeling unable to control things though this association was borderline significant (PR=1.73 (0.99, 3.00), p=0.05). There were no associations between water intermittency and other stress indicators (Table S9, Fig 3).

**Fig 3.**
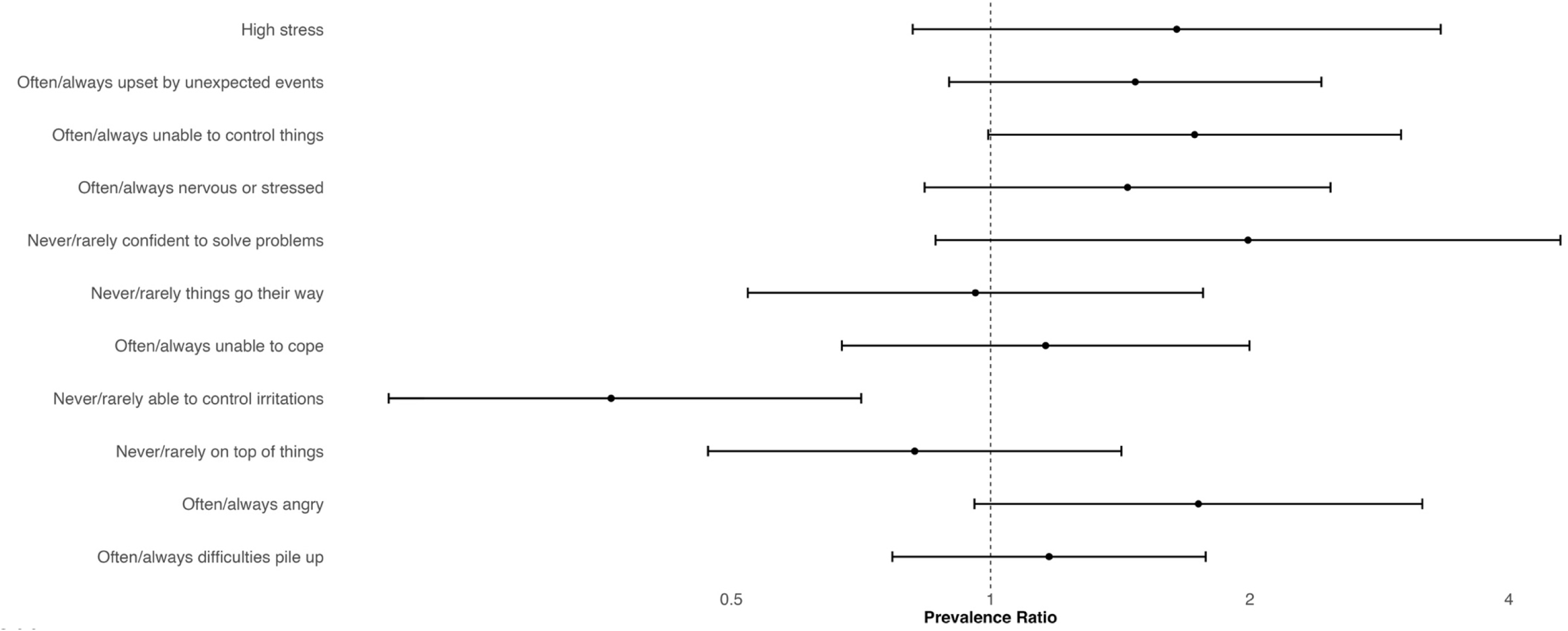
Forest plot of associations between water intermittency and self-reported caregiver stress. The circles denote point estimates for prevalence ratios and the horizontal lines denote the 95% confidence intervals. Models controlled for age of the respondent, age of the target child, total number of individuals living in household, respondent’s highest level of education, highest level of education level for anyone in household, asset-based household wealth quintile, average weekly expenditures by household, household food insecurity score, whether household used piped water for their primary water source, whether household’s designated handwashing station had soap, whether household had an improved latrine (as defined by the WHO), flooring material inside household, total number of animals in compound, and respondent’s report of the last time it rained.

When analyzed by categories of intermittency frequency, caregivers in households experiencing intermittency rarely had similar overall PSS scores, likelihood of experiencing high stress and likelihood of experiencing specific stress indicators compared to non-intermittent households (Table S10). Caregivers experiencing intermittency rarely were over 2 times more likely to report feeling angry (PR=2.21 (1.06, 4.61), p=0.04) but less likely to report feeling unable to control irritations (PR=0.30 (0.09, 0.94), p=0.039) than caregivers in non-intermittent households (Table S10). Caregivers experiencing intermittency rarely were also over 2.5 times more likely to report never/rarely feeling confident to solve problems though this association was borderline significant (PR=2.56 (0.95, 6.95), p=0.06); there were no other associations with stress indicators in this group after adjusting for confounders (Table S10). Caregivers in households experiencing intermittency sometimes, often, or always had higher PSS composite scores (ΔPSS=2.15 (0.24– 4.06), p=0.03) and were twice as likely to be experiencing high stress (PR=2.05, (0.99-4.25), p=0.05) compared to caregivers in non-intermittent households but the latter association was only borderline significant (Table S10). Caregivers in this group were also more likely to report feeling upset by unexpected events but less likely to report things not going their way and being unable to control irritations; these associations were only borderline significant (Table S10).

When analyzed by categories of intermittency duration, caregivers who experienced short intermittencies had higher PSS scores than non-intermittent households (ΔPSS=1.74 (0.08–3.40), p=0.04) (Table S11). Short intermittencies were also significantly associated with 2-3 times higher likelihood of caregivers feeling unable to control things (PR=1.95 (1.09, 3.50), p=0.03), nervous or stressed (2.12 (1.09, 4.13), p=0.03), not confident to solve problems (PR=3.39 (1.30, 8.81), p=0.01) and angry (PR=2.18 (1.07, 4.43), p=0.03) compared to non-intermittent households (Table S11). In contrast, caregivers in households experiencing longer intermittencies were less likely to report feeling unable to control irritations (PR=0.34 (0.13, 0.88), p=0.026) compared to non-intermittent households; there were no other associations between long water intermittencies and stress indicators.

The truncated household water insecurity (HWISE) score excludes the intermittency question to allow comparing scores between intermittent and non-intermittent households.

## 4. Discussion

In our study, approximately one third of households experienced at least one intermittency in the last four weeks, aligning with research from LMICs where intermittencies in water supply are a common challenge in urban and peri-urban settings (Adams et al., 2020; Aihara et al., 2015; Brewis et al., 2020; Nounkeu & Dharod, 2018; Workman et al., 2024). Households experiencing a water intermittency were more likely to de-prioritize hygiene practices, especially around animal management, and had higher water insecurity scores. They were also more likely to rely on stored drinking water and alternative water sources, but drinking water quality, as measured by *E. coli,* was not adversely affected by intermittencies. Children in households experiencing a water intermittency had higher prevalence of diarrhea, ARIs and ARIs with fever, while caregivers in these households reported significantly higher emotional stress levels, but primarily for short and frequent intermittencies. Our findings are broadly consistent with prior studies that have documented adverse effects of water intermittencies individually on water use behaviors, water quality, health or stress outcomes.

Prior research suggests that water access inequities often align with wealth disparities, with lower-income households disproportionately affected due to a reliance on communal sources (Adams et al., 2020; Jepson et al., 2017). In our study, households experiencing intermittencies were socioeconomically similar to those not experiencing intermittencies within the same geographic area. When assessing intermittency by sampling location, we found that 17% of all intermittencies occurred within a single geographic cluster, while the remaining intermittent households were spread across the other clusters. This suggests that water intermittency may not be solely driven by socioeconomic status but also by additional external geographic and infrastructural challenges, highlighting a more complex relationship between wealth and water intermittency (Adams et al., 2020).

Water intermittency is recognized as a component of water insecurity. Consistent with prior research, our study found that an intermittent water supply disrupts household water management and increases reliance on potentially unsafe water sources (Jeandron et al., 2015; Kumpel & Nelson, 2016). Notably, household water insecurity scores were low in our study (mean of 3.9 points out of a maximum possible of 36 points), and less than 10% (23) of households were classified as water insecure (HWISE score ≥12 points). Despite being relatively water secure, households that experienced at least one water intermittency in the last four weeks were substantially more likely to skip hygiene practices, such as bathing, laundry and handwashing after dirty activities. This supports previous studies linking water insecurity with reduced hygiene (Aihara et al., 2015; Ashraf et al., 2020; Freeman et al., 2014; Nounkeu & Dharod, 2018; Ross et al., 2023; Swarthout et al., 2020). Importantly, in our study, households experiencing an intermittency had similar reported handwashing during some key moments (after defecation, before eating or handling food or water) as non-intermittent households, consistent with evidence that washing hands after defecation is prioritized despite water limitations (Pickering et al., 2019; United Nations Children’s Fund (UNICEF) & World Health Organization (WHO), 2020). However, in our study, intermittent households were substantially less likely to report washing hands after handling animals and their feces or working outside (e.g. in the garden). Backyard animal husbandry is common in low- and middle-income countries and exposure to domestic animals is a risk factor for infectious diseases (Ercumen et al., 2017; Roess et al., 2013; Zambrano et al., 2014). Between 75-80% of households in our study reported having animals in the compound, and the number of animals per compound ranged from 6 to 9 (Table 1). Domestic soil is also increasingly recognized as a disease transmission pathway in settings where fecal waste is not isolated from the environment (Ercumen et al., 2018; Pickering et al., 2012). Our findings indicate that water conservation strategies associated with intermittencies may potentially increase zoonotic and soilborne exposure to enteric pathogens.

Intermittent households were more likely to obtain drinking water from secondary sources and use stored drinking water and less likely to use narrow-mouthed containers, which are more protective against contamination (Ercumen et al., 2015; Kumpel & Nelson, 2016). Increased reliance on stored water is a well-documented risk factor for microbial contamination due to prolonged storage and environmental exposure (Kumpel & Nelson, 2013). However, we did not find significant differences in *E. coli* prevalence or levels between intermittent and non-intermittent households. This finding contrasts with studies from other settings that reported a link between intermittency and microbial contamination of drinking water and suggests that, in our context, the increased risk of enteric and respiratory infections associated with water intermittencies may be more strongly driven by changes in hygiene-related behavior and limited water access and quantity than by differences in water quality.

It is possible that differences in water sources partly explain the lack of water quality effects from intermittencies in our study. While approximately 80% of both intermittent and non-intermittent households used an improved primary water source, households that experienced intermittencies were more likely to obtain their primary drinking water from a piped connection outside the compound (with a mean *E. coli* level of 81.3 log10-MPN/100 mL) and less likely to obtain it from a borehole (with a mean *E. coli* level of 217.3 log10-MPN/100 mL). While our analyses controlled for whether the household’s drinking water came from a piped source, our findings of no adverse water quality impacts from intermittencies may residually reflect the better quality of piped sources, which more commonly experienced intermittencies. Piped water interruptions in Blantyre are driven by a combination of infrastructural, operational, and financial challenges faced by the Blantyre Water Board (BWB). Aging infrastructure and widespread leakages account for almost 40% of physical water losses but demand regularly exceeds supply capacity, especially during dry periods (Kalulu & Hoko, 2010). High levels of non-revenue water—exceeding 50%—due to illegal connections, faulty meters, and system leaks further strain the utility’s resources (Song, 2022). Additionally, water access is highly unequal across zones, with peri-urban and elevated areas experiencing especially short and inconsistent supply due to low pressure and distribution issues (Kalulu & Hoko, 2010; Song, 2022). Our findings highlight a trade-off between water supply reliability and quality, with boreholes presenting a supply that is less likely to be interrupted but more likely to be contaminated than piped sources of household drinking water.

Children in intermittent households had significantly higher prevalence of diarrhea, ARI, and particularly ARI with fever. This finding strengthens existing evidence linking water intermittency to adverse health outcomes (Bivins et al., 2017; Ercumen et al., 2015; Rhue et al., 2023). Most research on water intermittencies and water insecurity has focused on diarrheal disease. A small number of studies have investigated relationships between water quality improvements and ARI (Ashraf et al., 2020; Kirby et al., 2019; Swarthout et al., 2020). Our findings suggest water intermittency may also increase ARI risk through pathways such as compromised hygiene, consistent with our findings of impaired handwashing practices in intermittent households. Emerging research has shown associations between water intermittencies and lessened ability to adhere to handwashing recommendations for COVID-19 prevention and higher risk of COVID-19 infections (Kumpel et al., 2022; Parisoto et al., 2024; Ray, 2020). Existing disease burden estimates for intermittent water supplies are focused on gastrointestinal infections (Bivins et al., 2017); our findings on ARIs indicate that these assessments may underestimate the total disease burden from water intermittencies. Additionally, when we analyzed health outcomes by intermittency frequency and duration, children in households with rare or short disruptions had significantly higher prevalence of caregiver-defined diarrhea while children with frequent or long disruptions had higher prevalence of ARIs, suggesting that short-term vs. long-term behavioral adaptations may differently impact enteric versus respiratory pathogen exposure. Further, almost one third of children in our study had used antibiotics at least once on the last four weeks, and infrequent and long intermittencies were associated with higher antibiotic use compared to non-intermittent households. Our findings suggest that the increased prevalence of enteric and respiratory infections among children experiencing intermittencies may translate to increased antibiotic use, highlighting antibiotic use and antimicrobial resistance as outcomes to investigate in future studies of water supply intermittencies.

A review conducted in 2016 found that predictable intermittencies, compared to irregular and unpredictable intermittencies, were the most disruptive in the lives of consumers across over 100 empirical case studies (Galaitsi et al., 2016). We found that shorter and more frequent intermittencies were associated with higher stress among caregivers, which may be due to greater uncertainty and need for adaptation strategies, supporting that even brief disruptions in water supply can have psychological effects. A recent study found that being able to predict a water intermittency reduced the stress response associated with it (Thomson et al., 2024). We were not able to test the influence of predictability on caregiver stress because most households who experienced an intermittency reported that they were not notified about it nor able to predict it. These findings align with prior research linking water insecurity to increased psychosocial distress, particularly among caregivers responsible for household water management (Adams, 2024; Jepson et al., 2017; Smiley & Stoler, 2020; Stevenson et al., 2012; Workman et al., 2022, 2024). Chronic stress, mental health and emotional wellbeing are important health dimensions to consider in the context of water insecurity. Our findings reinforce calls for integrating mental health considerations into water supply programs.

Climate change is expected to exacerbate water access challenges by increasing the frequency and severity of droughts, rainfall variability and extreme weather events. These stressors can prompt persistent water supply interruptions and disrupt traditional water sources, potentially increasing reliance on unprotected sources, as well as overcrowding and social conflict around water access. Additionally, climate change can alter seasonality, stressing households as protected, piped water may be less available during dry months, while unprotected sources such as surface water may be more accessible during the rainy season (Brewis et al., 2020; Guo et al., 2019). While our study did not assess the causes of the observed intermittencies, the water utilities report a complex set of causes, including compromised infrastructure, power shortages that disrupt pumping as well as water shortages and high demand during the dry season (Northern Region Water Board, 2025). As changing weather patterns, extreme events, and shifting seasonal cycles increasingly disrupt water availability, future research should quantify health and well-being impacts from increasingly unpredictable and intermittent water supplies (Cronk et al., 2024; Workman et al., 2024).

Our study integrated microbial, behavioral, health, and psychosocial dimensions of water intermittencies for a comprehensive assessment. This study also had limitations. Both water intermittencies and outcomes of interest were self-reported and may be subject to recall bias, potentially affecting our exposure and outcome classifications. Specifically, households experiencing intermittencies may be aware of health risks and therefore likely to overreport adverse health outcomes. However, our findings on our negative control outcome (rash) suggest that recall bias was unlikely to fully explain the associations observed. If recall bias were a major driver of our findings, we would expect to see similarly strong associations between water intermittency and the prevalence of children experiencing a rash, yet we did not observe such an association. We also note that while we selected rash as a negative control outcome under the assumption that it is causally independent of water intermittency, it is possible that hygiene behaviors or water-related stress could influence skin conditions. Further, the observational design of this study limits our ability to draw causal inferences between water intermittency and health/stress outcomes, and our findings may be affected by residual confounding from unaccounted-for differences between intermittent and non-intermittent households. Nevertheless, we recorded outcomes across the causal chain, including potential mediators, and conclude that our findings are internally consistent and biologically plausible because the observed direction of effects on mediating factors supports the observed associations with our study outcomes.

## 5. Conclusion

Our findings indicate that increased water intermittency is significantly associated with reduced hygiene practices, especially in the context of handling domestic animals, higher risk of respiratory infections and diarrhea in children, and increased psychological stress among caregivers. While our observational findings cannot determine causality, they suggest that improved household coping strategies— including safe water storage practices, use of safe alternative water sources during disruptions, low-flow hand hygiene strategies, water reuse for domestic hygiene, and advanced notification on upcoming disruptions—may help reduce health risks during water supply disruptions, with provision of an uninterrupted continuous water supply being the ultimate goal. Additionally, targeted support for caregiver stress may be beneficial, given the increasing evidence on the relationship between water insecurity and psychosocial outcomes. Future research should explore the long-term health consequences and implications of water intermittency, and explore interventions aimed at reducing the occurrence, frequency and duration of water supply intermittencies.

## Data Availability

All data produced in the present study are available upon reasonable request to the authors

## References

Adams, E. A. (2018). Thirsty slums in African cities: Household water insecurity in urban informal settlements of Lilongwe, Malawi. International Journal of Water Resources Development, 34(6), 869–887. 10.1080/07900627.2017.1322941

Adams, E. A. (2024). “Why Should a Married Man Fetch Water?” Masculinities, gender relations, and the embodied political ecology of urban water insecurity in Malawi. Social & Cultural Geography, 25(4), 582–600. 10.1080/14649365.2023.2183245

Adams, E. A., Stoler, J., & Adams, Y. (2020). Water insecurity and urban poverty in the Global South: Implications for health and human biology. American Journal of Human Biology, 32(1), e23368. 10.1002/ajhb.23368

Aihara, Y., Shrestha, S., Kazama, F., & Nishida, K. (2015). Validation of household water insecurity scale in urban Nepal. Water Policy, 17(6), 1019–1032. 10.2166/wp.2015.116

Arnold, B. F., & Ercumen, A. (2016). Negative Control Outcomes: A Tool to Detect Bias in Randomized Trials. JAMA, 316(24), 2597. 10.1001/jama.2016.17700

Ashraf, S., Islam, M., Unicomb, L., Rahman, M., Winch, P. J., Arnold, B. F., Benjamin-Chung, J., Ram, P. K., Colford, J. M., & Luby, S. P. (2020). Effect of Improved Water Quality, Sanitation, Hygiene and Nutrition Interventions on Respiratory Illness in Young Children in Rural Bangladesh: A Multi-Arm Cluster-Randomized Controlled Trial. The American Journal of Tropical Medicine and Hygiene, 102(5), 1124–1130. 10.4269/ajtmh.19-0769

Bivins, A. W., Sumner, T., Kumpel, E., Howard, G., Cumming, O., Ross, I., Nelson, K., & Brown, J. (2017). Estimating Infection Risks and the Global Burden of Diarrheal Disease Attributable to Intermittent Water Supply Using QMRA. Environmental Science & Technology, 51(13), 7542–7551. 10.1021/acs.est.7b01014

Brewis, A., DuBois, L. Z., Wutich, A., Adams, E. A., Dickin, S., Elliott, S. J., Empinotti, V. L., Harris, L. M., Ilboudo Nébié, E., & Korzenevica, M. (2024). Gender identities, water insecurity, and risk: Re theorizing the connections for a gender inclusive toolkit for water insecurity research. WIREs Water, 11(2), e1685. 10.1002/wat2.1685

Brewis, A., Workman, C., Wutich, A., Jepson, W., Young, S., & Household Water Insecurity Experiences – Research Coordination Network (HWISE RCN). (2020). Household water insecurity is strongly associated with food insecurity: Evidence from 27 sites in low and middle income countries. American Journal of Human Biology, 32(1), e23309. 10.1002/ajhb.23309

Burnham, J. P. (2021). Climate change and antibiotic resistance: A deadly combination. Therapeutic Advances in Infectious Disease, 8, 1–7. 10.1177/2049936121991374

Cohen, S., Kamarck, T., & Mermelstein, R. (1983). A Global Measure of Perceived Stress. Journal of Health and Social Behavior, 24(4), 385. 10.2307/2136404

Collins, S. M., Mbullo Owuor, P., Miller, J. D., Boateng, G. O., Wekesa, P., Onono, M., & Young, S. L. (2019). ‘I know how stressful it is to lack water!’ Exploring the lived experiences of household water insecurity among pregnant and postpartum women in western Kenya. Global Public Health, 14(5), 649–662. 10.1080/17441692.2018.1521861

Coulibaly, J. Y., Mbow, C., Sileshi, G. W., Beedy, T., Kundhlande, G., & Musau, J. (2015). Mapping Vulnerability to Climate Change in Malawi: Spatial and Social Differentiation in the Shire River Basin. American Journal of Climate Change, 4(3), Article 3. 10.4236/ajcc.2015.43023

Cronk, R., Tracy, J. W., & Bartram, J. (2024). The influence of seasonality and multiple water source use on household water service levels. Cleaner Water, 1, 100012. 10.1016/j.clwat.2024.100012

Ercumen, A., Arnold, B. F., Kumpel, E., Burt, Z., Ray, I., Nelson, K., & Colford, J. M. (2015). Upgrading a Piped Water Supply from Intermittent to Continuous Delivery and Association with Waterborne Illness: A Matched Cohort Study in Urban India. PLOS Medicine, 12(10), e1001892. 10.1371/journal.pmed.1001892

Ercumen, A., Naser, A. Mohd., Unicomb, L., Arnold, B. F., Colford Jr., J. M., & Luby, S. P. (2015). Effects of Source- versus Household Contamination of Tubewell Water on Child Diarrhea in Rural Bangladesh: A Randomized Controlled Trial. PLOS ONE, 10(3), e0121907.

Ercumen, A., Pickering, A. J., Kwong, L. H., Arnold, B. F., Parvez, S. M., Alam, M., Sen, D., Islam, S., Kullmann, C., Chase, C., Ahmed, R., Unicomb, L., Luby, S. P., & Colford, J. M. Jr. (2017). Animal Feces Contribute to Domestic Fecal Contamination: Evidence from E. coli Measured in Water, Hands, Food, Flies, and Soil in Bangladesh. Environmental Science & Technology, 51(15), 8725–8734. 10.1021/acs.est.7b01710

Ercumen, A., Pickering, A. J., Kwong, L. H., Mertens, A., Arnold, B. F., Benjamin-Chung, J., Hubbard, A. E., Alam, M., Sen, D., Islam, S., Rahman, Md. Z., Kullmann, C., Chase, C., Ahmed, R., Parvez, S. M., Unicomb, L., Rahman, M., Ram, P. K., Clasen, T., … Colford, J. M. (2018). Do Sanitation Improvements Reduce Fecal Contamination of Water, Hands, Food, Soil, and Flies? Evidence from a Cluster-Randomized Controlled Trial in Rural Bangladesh. Environmental Science & Technology, 52(21), 12089–12097. 10.1021/acs.est.8b02988

Ferrante, M., Rogers, D., Mugabi, J., & Casinini, F. (2024). A Laboratory Investigation on the Effects of Intermittent Water Supply and Remedial Measures. Water Resources Research, 60(2), e2023WR035282. 10.1029/2023WR035282

*Find Point Clusters—ArcGIS Online | Documentation*. (n.d.). Retrieved February 21, 2025, from https://doc.arcgis.com/en/arcgis-online/analyze/find-point-clusters-mv.htm#LI_6ABE0F9A58C441DE9FD578CE227B4476

Freeman, M. C., Stocks, M. E., Cumming, O., Jeandron, A., Higgins, J. P. T., Wolf, J., Prüss Ustün, A., Bonjour, S., Hunter, P. R., Fewtrell, L., & Curtis, V. (2014). Systematic review: Hygiene and health: systematic review of handwashing practices worldwide and update of health effects. Tropical Medicine & International Health, 19(8), 906–916. 10.1111/tmi.12339

Galaitsi, S., Russell, R., Bishara, A., Durant, J., Bogle, J., & Huber-Lee, A. (2016). Intermittent Domestic Water Supply: A Critical Review and Analysis of Causal-Consequential Pathways. Water, 8(7), 274. 10.3390/w8070274

Greenwood, E. E., Lauber, T., Van Den Hoogen, J., Donmez, A., Bain, R. E. S., Johnston, R., Crowther, T. W., & Julian, T. R. (2024). Mapping safe drinking water use in low- and middle-income countries. Science, 385(6710), 784–790. 10.1126/science.adh9578

Guo, D., Thomas, J., Lazaro, A., Mahundo, C., Lwetoijera, D., Mrimi, E., Matwewe, F., & Johnson, F. (2019). Understanding the impacts of short term climate variability on drinking water source quality: Observations from three distinct climatic regions in Tanzania. GeoHealth, 3(4), 84–103. 10.1029/2018GH000180

Hanif, S., Momo, J.-E.-T., Jahan, F., Goldberg, L., Herbert, N., Yeamin, A., Shoab, A. K., Akhter, R. M., Roy, S. K., Heitmann, G. B., Ercumen, A., Rahman, M., Tofail, F., Wong-Parodi, G., & Benjamin-Chung, J. (2024). Flooding and elevated prenatal depression in a climate-sensitive community in rural Bangladesh: A mixed methods study. medRxiv, 2024.11.25.24317922. 10.1101/2024.11.25.24317922

Harawa, M. M., Hoko, Z., Misi, S., & Maliano, S. (2016). Investigating the management of unaccounted for water for Lilongwe Water Board, Malawi. Journal of Water, Sanitation and Hygiene for Development, 6(3), 362–376. 10.2166/washdev.2016.013

Harris, K. M., Gaffey, A. E., Schwartz, J. E., Krantz, D. S., & Burg, M. M. (2023). The Perceived Stress Scale as a Measure of Stress: Decomposing Score Variance in Longitudinal Behavioral Medicine Studies. Annals of Behavioral Medicine, 57(10), 846–854. 10.1093/abm/kaad015

Hassan, M. Z., Monjur, M. R., Biswas, M. A. A. J., Chowdhury, F., Kafi, M. A. H., Braithwaite, J., Jaffe, A., & Homaira, N. (2021). Antibiotic use for acute respiratory infections among under-5 children in Bangladesh: A population-based survey. BMJ Global Health, 6(4), e004010. 10.1136/bmjgh-2020-004010

Hayward, C., Ross, K. E., Brown, M. H., & Whiley, H. (2020). Water as a Source of Antimicrobial Resistance and Healthcare-Associated Infections. Pathogens, 9(8), 667. 10.3390/pathogens9080667

Holm, R. H., Tembo, M., Kasulo, V., Gavanala, M. B., & Chilongo, L. (2022). Institutional, technical and financial sustainability of rural piped drinking water supply on a freshwater island: Case study of Likoma Island, Malawi. *Lakes & Reservoirs: Science*, Policy and Management for Sustainable Use, 27(2), e12403. 10.1111/lre.12403

Hornsby, G., D. Ibitoye, T., Keelara, S., & Harris, A. (2023). Validation of a modified IDEXX defined-substrate assay for detection of antimicrobial resistant E. coli in environmental reservoirs. Environmental Science: Processes & Impacts, 25(1), 37–43. 10.1039/D2EM00189F

Jeandron, A., Saidi, J. M., Kapama, A., Burhole, M., Birembano, F., Vandevelde, T., Gasparrini, A., Armstrong, B., Cairncross, S., & Ensink, J. H. J. (2015). Water Supply Interruptions and Suspected Cholera Incidence: A Time-Series Regression in the Democratic Republic of the Congo. PLOS Medicine, 12(10), e1001893. 10.1371/journal.pmed.1001893

Jepson, W. E., Wutich, A., Colllins, S. M., Boateng, G. O., & Young, S. L. (2017). Progress in household water insecurity metrics: A cross-disciplinary approach. WIREs Water, 4(3), e1214. 10.1002/wat2.1214

Kalulu, K., & Hoko, Z. (2010). Assessment of the performance of a public water utility: A case study of Blantyre Water Board in Malawi. *Physics and Chemistry of the Earth*, Parts A/B/C, 35(13), 806–810. 10.1016/j.pce.2010.07.017

Kirby, M. A., Nagel, C. L., Rosa, G., Zambrano, L. D., Musafiri, S., Ngirabega, J. de D., Thomas, E. A., & Clasen, T. (2019). Effects of a large-scale distribution of water filters and natural draft rocket-style cookstoves on diarrhea and acute respiratory infection: A cluster-randomized controlled trial in Western Province, Rwanda. PLOS Medicine, 16(6), e1002812. 10.1371/journal.pmed.1002812

Kumpel, E., Billava, N., Nayak, N., & Ercumen, A. (2022). Water use behaviors and water access in intermittent and continuous water supply areas during the COVID-19 pandemic. Journal of Water and Health, 20(1), 139–148. 10.2166/wh.2021.184

Kumpel, E., & Nelson, K. L. (2013). Comparing microbial water quality in an intermittent and continuous piped water supply. Water Research, 47(14), 5176–5188. 10.1016/j.watres.2013.05.058

Kumpel, E., & Nelson, K. L. (2016). Intermittent Water Supply: Prevalence, Practice, and Microbial Water Quality. Environmental Science & Technology, 50(2), 542–553. 10.1021/acs.est.5b03973

Lipsitch, M., Tchetgen Tchetgen, E., & Cohen, T. (2010). Negative Controls: A Tool for Detecting Confounding and Bias in Observational Studies. Epidemiology, 21(3), 383. 10.1097/EDE.0b013e3181d61eeb

MacAllister, D. J., Nedaw, D., Kebede, S., Mkandawire, T., Makuluni, P., Shaba, C., Okullo, J., Owor, M., Carter, R., Chilton, J., Casey, V., Fallas, H., & MacDonald, A. M. (2022). Contribution of physical factors to handpump borehole functionality in Africa. Science of The Total Environment, 851, 158343. 10.1016/j.scitotenv.2022.158343

McCarthy, N., Kilic, T., Brubaker, J., Murray, S., & Fuente, A. de la. (2021). Droughts and floods in Malawi: Impacts on crop production and the performance of sustainable land management practices under weather extremes. Environment and Development Economics, 26(5–6), 432–449. 10.1017/S1355770X20000455

Nounkeu, C. D., & Dharod, J. M. (2018). Water insecurity among rural households of West Cameroon: Lessons learned from the field. *Journal of Water*, Sanitation and Hygiene for Development, 8(3), 585–594. 10.2166/washdev.2018.148

O’Brien, L. A., Snyder, J. S., Garn, J. V., Kann, R., Júnior, A., McGunegill, S., Muneme, B., Manuel, J. L., Nalá, R., Levy, K., & Freeman, M. C. (2024). Water, food, and mental well-being: Associations between drinking water source, household water and food insecurity, and mental well-being of low-income pregnant women in urban Mozambique. PLOS Water, 3(6), e0000219. 10.1371/journal.pwat.0000219

Parisoto, N. M. S. F., Lopes, L. C. P., Inokuma, A. A., Santos, E. A. D., Pavani, N. P. G., Santos, P. S. S., & Bastos, R. S. (2024). Intermittent Water Supply, Primary Health Coverage, and COVID-19 Among Older Adults: Addressing Structural Inequities in a Medium-Sized Brazilian City. 10.22541/au.173542779.91364444/v1

Pickering, A. J., Ercumen, A., Arnold, B. F., Kwong, L. H., Parvez, S. M., Alam, M., Sen, D., Islam, S., Kullmann, C., Chase, C., Ahmed, R., Unicomb, L., Colford, J. M. Jr., & Luby, S. P. (2018). Fecal Indicator Bacteria along Multiple Environmental Transmission Pathways (Water, Hands, Food, Soil, Flies) and Subsequent Child Diarrhea in Rural Bangladesh. Environmental Science & Technology, 52(14), 7928–7936. 10.1021/acs.est.8b00928

Pickering, A. J., Julian, T. R., Marks, S. J., Mattioli, M. C., Boehm, A. B., Schwab, K. J., & Davis, J. (2012). Fecal Contamination and Diarrheal Pathogens on Surfaces and in Soils among Tanzanian Households with and without Improved Sanitation. Environmental Science & Technology, 46(11), 5736–5743. 10.1021/es300022c

Pickering, A. J., Njenga, S. M., Steinbaum, L., Swarthout, J., Lin, A., Arnold, B. F., Stewart, C. P., Dentz, H. N., Mureithi, M., Chieng, B., Wolfe, M., Mahoney, R., Kihara, J., Byrd, K., Rao, G., Meerkerk, T., Cheruiyot, P., Papaiakovou, M., Pilotte, N., … Null, C. (2019). Effects of single and integrated water, sanitation, handwashing, and nutrition interventions on child soil-transmitted helminth and Giardia infections: A cluster-randomized controlled trial in rural Kenya. PLOS Medicine, 16(6), e1002841. 10.1371/journal.pmed.1002841

Pieh, C., Dale, R., Plener, P. L., Humer, E., & Probst, T. (2022). Stress levels in high-school students after a semester of home-schooling. European Child & Adolescent Psychiatry, 31(11), 1847–1849. 10.1007/s00787-021-01826-2

Ray, I. (2020). Viewpoint – Handwashing and COVID-19: Simple, right there…? World Development, 135, 105086. 10.1016/j.worlddev.2020.105086

Rayasam, S. D. G., Ray, I., Smith, K. R., & Riley, L. W. (2019). Extraintestinal Pathogenic Escherichia coli and Antimicrobial Drug Resistance in a Maharashtrian Drinking Water System. The American Journal of Tropical Medicine and Hygiene, 100(5), 1101–1104. 10.4269/ajtmh.18-0542

Rhee, C., Aol, G., Ouma, A., Audi, A., Muema, S., Auko, J., Omore, R., Odongo, G., Wiegand, R. E., Montgomery, J. M., Widdowson, M.-A., O’Reilly, C. E., Bigogo, G., & Verani, J. R. (2019). Inappropriate use of antibiotics for childhood diarrhea case management— Kenya, 2009–2016. BMC Public Health, 19(S3), 468. 10.1186/s12889-019-6771-8

Rhue, S. J., Torrico, G., Amuzie, C., Collins, S. M., Lemaitre, A., Workman, C. L., Rosinger, A. Y., Pearson, A. L., Piperata, B. A., Wutich, A., Brewis, A., & Stoler, J. (2023). The effects of household water insecurity on child health and well-being. WIREs Water, 10(6), e1666. 10.1002/wat2.1666

Roess, A. A., Winch, P. J., Ali, N. A., Akhter, A., Afroz, D., El Arifeen, S., Darmstadt, G. L., Baqui, A. H., &. (2013). Animal Husbandry Practices in Rural Bangladesh: Potential Risk Factors for Antimicrobial Drug Resistance and Emerging Diseases. The American Society of Tropical Medicine and Hygiene, 89(5), 965–970. 10.4269/ajtmh.12-0713

Ross, I., Bick, S., Ayieko, P., Dreibelbis, R., Wolf, J., Freeman, M. C., Allen, E., Brauer, M., & Cumming, O. (2023). Effectiveness of handwashing with soap for preventing acute respiratory infections in low-income and middle-income countries: A systematic review and meta-analysis. The Lancet, 401(10389), 1681–1690. 10.1016/S0140-6736(23)00021-1

Smiley, S. L., & Stoler, J. (2020). Socio-environmental confounders of safe water interventions. WIREs Water, 7(3), e1438. 10.1002/wat2.1438

Song, C. (2022). Analysis of the organization, management and operation of Blantyre Water Board in Malawi: Future reforms and perspectives. Environmental Quality Management, 32(2), 287–294. 10.1002/tqem.21843

Stevenson, E. G. J., Greene, L. E., Maes, K. C., Ambelu, A., Tesfaye, Y. A., Rheingans, R., & Hadley, C. (2012). Water insecurity in 3 dimensions: An anthropological perspective on water and women’s psychosocial distress in Ethiopia. Social Science & Medicine, 75(2), 392–400. 10.1016/j.socscimed.2012.03.022

Swarthout, J., Ram, P. K., Arnold, C. D., Dentz, H. N., Arnold, B. F., Kalungu, S., Lin, A., Njenga, S. M., Stewart, C. P., Colford, J. M., Null, C., & Pickering, A. J. (2020). Effects of Individual and Combined Water, Sanitation, Handwashing, and Nutritional Interventions on Child Respiratory Infections in Rural Kenya: A Cluster-Randomized Controlled Trial. The American Journal of Tropical Medicine and Hygiene, 102(6), 1286– 1295. 10.4269/ajtmh.19-0779

Taviani, E., Van Den Berg, H., Nhassengo, F., Nguluve, E., Paulo, J., Pedro, O., & Ferrero, G. (2022). Occurrence of waterborne pathogens and antibiotic resistance in water supply systems in a small town in Mozambique. BMC Microbiology, 22(1), 243. 10.1186/s12866-022-02654-3

Tekleab, A. M., Asfaw, Y., Weldetsadik, A., & Amaru, G. M. (2017). Antibiotic prescribing practice in the management of cough or diarrhea among children attending hospitals in Addis Ababa: A cross-sectional study. Pediatric Health, Medicine and Therapeutics, Volume 8, 93–98. 10.2147/PHMT.S144796

Thomson, P., Pearson, A. L., Kumpel, E., Guzmán, D. B., Workman, C. L., Fuente, D., Wutich, A., Stoler, J., & Household Water Insecurity Experiences Research Coordination Network (HWISE-RCN). (2024). Water Supply Interruptions Are Associated with More Frequent Stressful Behaviors and Emotions but Mitigated by Predictability: A Multisite Study. Environmental Science & Technology, 58(16), 7010–7019. 10.1021/acs.est.3c08443

Tokajian, S., & Hashwa, F. (2004). Phenotypic and genotypic identification of Aeromonas spp. Isolated from a chlorinated intermittent water distribution system in Lebanon. Journal of Water and Health, 2(2), 115–122. 10.2166/wh.2004.0011

United Nations Children’s Fund (UNICEF), & World Health Organization (WHO). (2020). State of the world’s sanitation: An urgent call to transform sanitation for better health, environments, economies and societies. United Nations Children’s Fund (UNICEF) and the World Health Organization. https://www.unicef.org/reports/state-worlds-sanitation-2020

United Nations Children’s Fund (UNICEF), & World Health Organization (WHO). (2023). *Progress on household drinking water, sanitation and hygiene 2000–2022: Special focus on gender*.

United Nations, U. (2025). Human Development Reports: Country Insights. In Human Development Reports. United Nations. https://hdr.undp.org/data-center/country-insights

WHO. (2021). WHO Integrated Global Surveillance on ESBL-Producing E. Coli Using a One Health Approach: Implementation and Opportunities. World Health Organization.

WHO. (2023, November 13). Drinking-water. World Health Organization. http://who.int/news-room/fact-sheets/detail/drinking-water

Workman, C. L., Miller, J. D., Shah, S. H., Maes, K., Tesfaye, Y., & Mapunda, K. M. (2024). Frequency and perceived difficulty of household water experiences in Morogoro, Tanzania: Evidence of the psychosocial burden of water insecurity. SSM - Mental Health, 5, 100295. 10.1016/j.ssmmh.2023.100295

Workman, C. L., Stoler, J., Harris, A., Ercumen, A., Kearns, J., & Mapunda, K. M. (2022). Food, water, and sanitation insecurities: Complex linkages and implications for achieving WASH security. Global Public Health, 17(11), 3060–3075. 10.1080/17441692.2021.1971735

Workman, C. L., & Ureksoy, H. (2017). Water insecurity in a syndemic context: Understanding the psycho-emotional stress of water insecurity in Lesotho, Africa. Social Science & Medicine, 179, 52–60. 10.1016/j.socscimed.2017.02.026

Wutich, A., Brewis, A., & Tsai, A. (2020). Water and mental health. WIREs Water, 7(5), e1461. 10.1002/wat2.1461

Young, S. L., Boateng, G. O., Jamaluddine, Z., Miller, J. D., Frongillo, E. A., Neilands, T. B., Collins, S. M., Wutich, A., Jepson, W. E., & Stoler, J. (2019). The Household Water InSecurity Experiences (HWISE) Scale: Development and validation of a household water insecurity measure for low-income and middle-income countries. BMJ Global Health, 4(5), e001750. 10.1136/bmjgh-2019-001750

Yu, D., Ryu, K., Zhi, S., Otto, S. J. G., & Neumann, N. F. (2022). Naturalized Escherichia coli in Wastewater and the Co-evolution of Bacterial Resistance to Water Treatment and Antibiotics. Frontiers in Microbiology, 13. 10.3389/fmicb.2022.810312

Zambrano, L. D., Levy, K., Menezes, N. P., & Freeman, M. C. (2014). Human diarrhea infections associated with domestic animal husbandry: A systematic review and meta-analysis. Transactions of The Royal Society of Tropical Medicine and Hygiene, 108(6), 313–325. 10.1093/trstmh/tru056

