## Supplementary Materials for "Associations between water supply intermittencies and drinking water quality, child health, and caregiver emotional stress in peri-urban Malawi"

**Table of Contents**

**Text S1.** Adjusted model covariates ____________________________________________Page 1

**Table S1.** Household characteristics by intermittency frequency and duration __________ Page 2

**Table S2**. Water quality according to water source type ____________________________ Page 3

**Table S3.** Water quality vs. binary intermittency (N=236) __________________________Page 4

**Table S4.** Water quality vs. categorical intermittency frequency (N=236) ______________Page 4

**Table S5.** Water quality vs. categorical intermittency duration (N=232)________________Page 4

**Table S6.** Child health outcomes vs. binary intermittency (N=292) ___________________Page 5

**Table S7.** Child health outcomes vs. categorical intermittency frequency (N=292) _______Page 5

**Table S8.** Child health outcomes vs. categorical intermittency duration (N=287)_________Page 5

**Table S9.** Caregiver stress vs. binary intermittency (N=237) ________________________Page 6

**Table S10.** Caregiver stress vs. categorical intermittency frequency (N=237)___________ Page 7

**Table S11.** Caregiver stress vs. categorical intermittency duration (N=233) ____________ Page 7

**Text S1.** Adjusted model covariates

All models controlled for:

Household's primary water source piped

Household has an improved latrine

Household has handwashing station with soap (observed)

Number of animals living on the compound

Age of respondent

Total number of people living in the household

Respondent's highest level of education

Highest education level in the household

Household wealth index quintile

Amount of money spent per week by the household, in USD

Floor material in the household

Respondent's report of last time it rained

Models for water quality outcomes additionally controlled for:

Water sample obtained from piped source

Water sample reported to be treated

Models for health and stress outcomes additionally controlled for:

Child age, in months

Food insecurity index (HFIAS score)

We also considered the following adjustment covariates but did not control for them because they had <5% variation in the study sample (i.e. one category dominated across all households): Roof material, wall material, and for respiratory outcomes, fuel type, stove type and presence of windows in the dwelling as predictors of indoor air quality.

**Table S1.** Hygiene practices, water storage and secondary water sources by intermittency frequency and duration

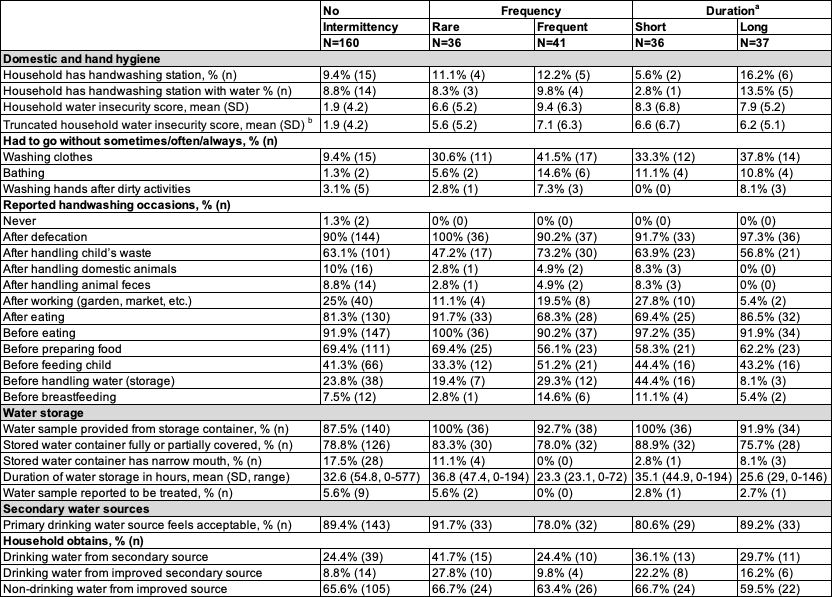

^a^ 4 households dropped from analysis because of missing data on intermittency duration.

^b^ The truncated household water insecurity (HWISE) score excludes the intermittency question to allow comparing scores between intermittent and non-intermittent households.

**Table S2**. Prevalence and most probable number (MPN) of *E. coli* and cefotaxime-resistant *E. coli* by water source type

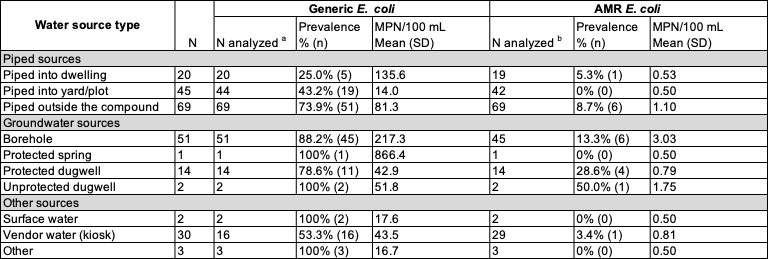

AMR: Antimicrobial-resistant, SD: Standard deviation, log-10 MPN/100 mL: log10-transformed most probable number per 100 mL

^a^ 1 household missing water sample

^b^ 10 water samples not processed for AMR *E. coli*

**Table S3**. Water quality vs. binary intermittency (N=236)

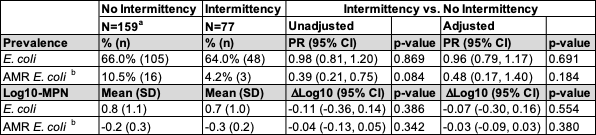

AMR: Antimicrobial-resistant, MPN: Most probable number, SD: Standard deviation, PR: Prevalence ratio, CI: Confidence interval, ΔLog10: Difference in log10-transformed most probable number

^a^ 1 household missing water sample

^b^ 10 water samples not processed for AMR *E. coli*

**Table S4**. Water quality vs. categorical intermittency frequency (N=236)

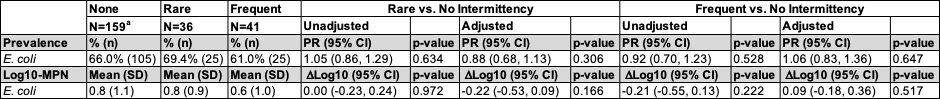

AMR: Antimicrobial-resistant, MPN: Most probable number, SD: Standard deviation, PR: Prevalence ratio, CI: Confidence interval, ΔLog10: Difference in log10-transformed most probable number

^a^ 1 household missing water sample

**Table S5**. Water quality vs. categorical intermittency duration (N=232^a^)

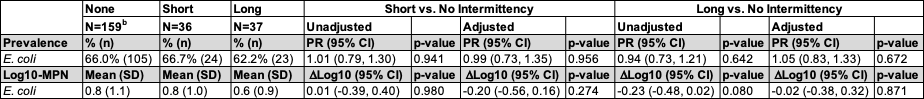

AMR: Antimicrobial-resistant, MPN: Most probable number, SD: Standard deviation, PR: Prevalence ratio, CI: Confidence interval, ΔLog10: Difference in log10-transformed most probable number

^a^ 4 households dropped from analysis because of missing data on intermittency duration

^b^ 1 household missing water sample

**Table S6**. Child health outcomes vs. binary intermittency (N=292)

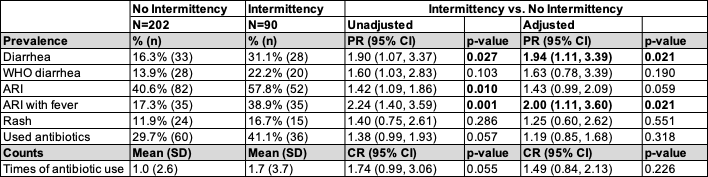

SD: Standard deviation, PR: Prevalence ratio, CI: Confidence interval, CR: Count ratio

**Table S7**. Child health outcomes vs. categorical intermittency frequency (N=292)

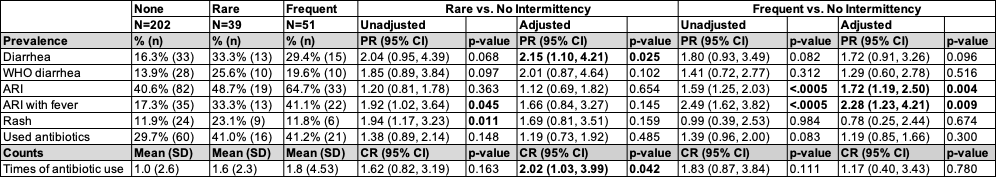

SD: Standard deviation, PR: Prevalence ratio, CI: Confidence interval, CR: Count ratio

**Table S8**. Child health outcomes vs. categorical intermittency duration (N=287^a^)

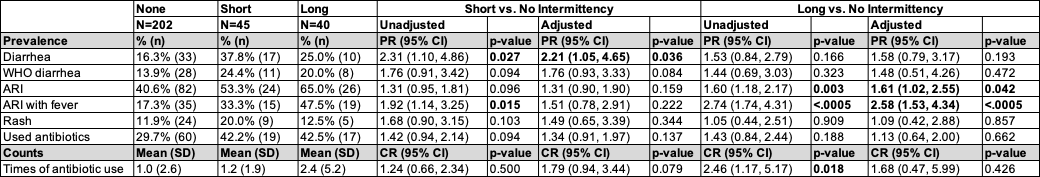

SD: Standard deviation, PR: Prevalence ratio, CI: Confidence interval, CR: Count ratio

^a^ 5 children dropped from analysis because of missing data on intermittency duration

**Table S9**. Caregiver stress vs. binary intermittency (N=237)

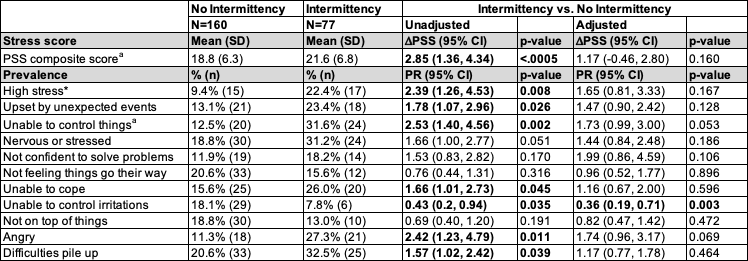

PSS: Perceived stress scale, SD: Standard deviation, ΔPSS: Difference in PSS composite score, CI: Confidence interval, PR: Prevalence ratio

^a^ PSS composite score could not be calculated for 1 household that had a missing response to one question in the 10-question scale

**Table S10**. Caregiver stress vs. categorical intermittency frequency (N=237)

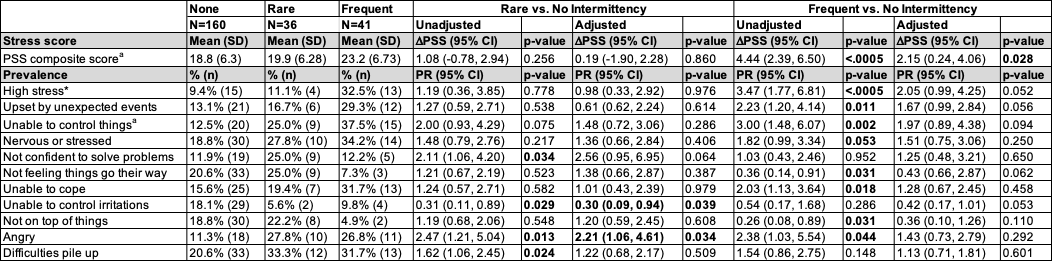

PSS: Perceived stress scale, SD: Standard deviation, ΔPSS: Difference in PSS composite score, CI: Confidence interval, PR: Prevalence ratio

^a^ PSS composite score could not be calculated for 1 household that had a missing response to one question in the 10-question scale

**Table S11**. Caregiver stress vs. categorical intermittency duration (N=233^a^)

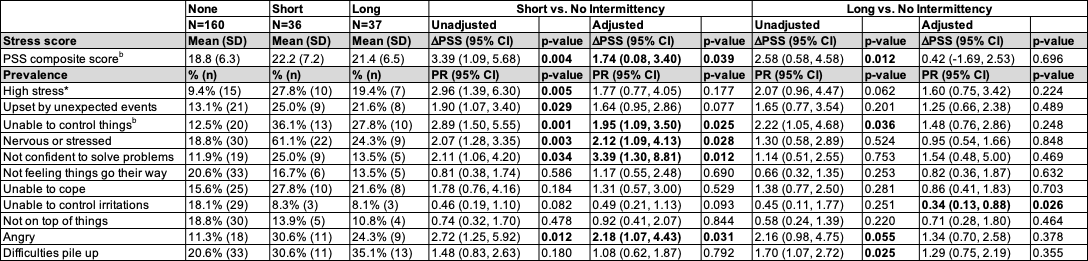

PSS: Perceived stress scale, SD: Standard deviation, ΔPSS: Difference in PSS composite score, CI: Confidence interval, PR: Prevalence ratio

^a^ 4 households dropped from analysis because of missing data on intermittency duration

^b^ PSS composite score could not be calculated for 1 household that had a missing response to one question in the 10-question scale
